# Mimicking cancer therapy in an agent-based model: the case of hepatoblastoma

**DOI:** 10.1101/2025.02.07.25321671

**Authors:** Alessandro Ravoni, Enrico Mastrostefano, Roland Kappler, Carolina Armengol, Filippo Castiglione, Christine Nardini

## Abstract

Hepatoblastoma is the most common pediatric liver cancer and represents a serious clinical challenge as no effective therapies have yet been found for advanced states and relapses of the disease. In this work, we use a well-established agent-based model of the immune response now equipped with anti-cancer therapy response to study the evolution of the disease and the role of the immune system in its containment, in particular by simulating the course of hepatoblastoma over three years in a population of virtual patients with mortality and symptom onset rates mimicking clinical ones. These results, combined with the ability to observe the dynamics of cellular entities at the microscopic scale and the key chemical signals involved in disease progression, make the model a valuable resource for future research on *in silico* trials.

## 1. Introduction

Hepatoblastoma (HB) is the most common liver cancer in pediatric patients accounting for 80% of hepatic tumors in children [1, 2, 3]. The standard therapy for HB, consisting of chemotherapy and surgery, remains ineffective in relapses and advanced states of the disease, and continuous efforts are therefore made to develop new therapeutic strategies and improve prognosis [4, 5, 6].

A multidisciplinary approach involving clinical expertise and *in silico* models may be of great help in this endeavor and notably for testing and development of new therapies. In this perspective, we here present the study of the combined effect of the activity of the immune system (IS) elicited by standard cancer treatment (drug therapy plus resection) against HB by using an agent-based model (ABM) of the immune response, namely C-IMMSIM [7, 8, 9, 10]. C-IMMSIM simulates the dynamics of computational entities (*agents*) representing cells and molecules participating in the adaptive response against one or more antigens. In Castiglione et al. [11] C-IMMSIM has already been applied to the study of a generic solid cancer by mimicking the immune response elicited by the injection of a vaccine. In this work, we extend the features of the ABM to represent cancer cells and their interactions with other cells and molecules that participate in the immune response. Further, we introduce the effect of chemotherapy as a cell death process occurring with a given probability, with parameters regulating the death process being estimated from experimental data.

Our goal is to develop a simulator that models the disease progression of HB in both untreated and treated patients, using data from the literature and the iPC Consortium (iPC - Individualized PaediatricCure, ref. 826121, H2020-SC1-DTH-2018-1).

To achieve this -after careful calibration of the simulatorwe mimic the progression of a large cohort of virtual patients, from diagnosis through therapy up to the final outcome. Each virtual patient is initialized with a variable number of cancer cells and an individual immune repertoire and is further randomly assigned to a diagnostic group. As the tumor volume grows and surpasses a predefined threshold, symptoms appear, triggering the start of therapy. Treatment consists of an initial period of chemotherapy, followed by surgical resection, and then additional chemotherapy. Immunological variables -such as immune cells abundances and cytokines concentrations-as well as measurable features of tumor progression (e.g. cell count or tumor volume) represent the disease trajectory and are monitored throughout the entire experiment and for three years after the completion of treatment.

The paper proceeds from the description of C-IMMSIM in Section 2, with details on all features added to mimic cancers and specifically HB, to the results obtained in simulating HB progression in Section 3, overall discussed in the concluding Section 4.

## 2. Models and methods

We first provide in Section 2.1 a general description of C-IMMSIM, a thoroughly evaluated tool supported by numerous publications [7, 8, 9, 10]. We then introduce the novelties of this work, which include: in Section 2.2 the cancer growth model and the cancer-IS interactions; in Section 2.3 the adaptation to the specific case of HB; and finally the procedure to calibrate the model parameters in Section 2.4.

### 2.1. The model

C-IMMSIM consists of stochastic cellular automata evolving in a three-dimensional lattice (representing several cubic millimeters of a lymph node of a vertebrate animal or milliliters of peripheral blood). Such automata can be of two types: molecules and cells.

*Molecules* are the simplest simulated entities, mainly characterised by their number, position (in the 3-D lattice) and binding site. Examples of simulated molecules are antigens (Ag, like bacteria and generic viruses or tumor-associated antigens, TAA), antibodies (AB), immune complexes, or intercellular signaling molecules (cytokines and danger signals).

*Cells* are Stochastic Finite State Machines, whose state changes are represented by stochastic events that depend on interactions with other entities. Each cell is spatially located in the model and labelled with a state variable that determines its behaviour and interactions with other entities (e.g., an *inactive* lymphocytes can switch to the *active* state) following interaction with an antigen-presenting cell. C-IMMSIM takes into account the dynamics of cells such as lymphocytes B (B), lymphocytes T helper (TH), lymphocytes T killer or cytotoxic (TC), macrophages (MA), epithelial cells (EP), plasma lymphocytes B (PLB) and natural killer cells (NK).

Entities (molecules and cells) *e* are associated with *N*_*e*_ binary strings representing their binding sites. In this framework, the different lymphocyte receptors’ subtypes - that constitute an individual’s immune repertoire-are represented by a set of binary strings. Affinity between two entities is hence efficiently represented as the Hamming distance [12] between such binary strings. Note that for some entities, such as cytokines, *N*_*e*_ = 0.

During the simulation, which proceeds in discrete time steps [13], the entities move across the lattice and interact with other entities, provided they are simultaneously at the same lattice coordinates, with the rules that implement the interactions being executed in a randomized order.

Immune cell migration is itself a stochastic process: at each time step, cells can move freely in all directions, at most by one unit in the lattice and with a probability dependent on the cellular density at that coordinates. Details on the movement of agents and their interactions in C-IMMSIM can be found in Castiglione and Celada [10].

Finally, it is worth noting that, given the stochastic nature of the simulations, fixed values of the model parameters produce different realisations of the system dynamics. In particular, the simulation runs differ from each other in the immune repertoire and in the initial number of immune cells -i.e., the immunological initial state-as well as in the occurrence of probabilistic events during the course of the simulation. In this sense, each simulation run can be associated with a specific virtual patient. By simulating several virtual patients, we can mimic the evolution of the disease within a population of individuals whose immune response entities’ dynamics are reproduced quantitatively.

### 2.2. Adding cancer cells to the model

Cancer development is represented by adding a new Stochastic Finite State Machine that simulates cancer cells and the mechanisms that affect their population. The changes in cell numbers over time *N*(*t*) are governed by stochastic processes that depend on the state of the system at any given time. These changes are modeled through four primary mechanisms: (i) cancer growth through cell duplication, leading to an increase in cell numbers; (ii) the IS interaction with cancer cells, which reduces their number; (iii) the impact of drugs and (iv) surgical removal of cancer cells. In the following we details each of these four processes in a dedicated subsection.

#### (i) Cancer growth

We model cancer growth as a density- and space-constrained replication process occurring at the single-cell level, i.e., at each time step, we compute the offspring generated by each cancer cell in the system. A detailed description of the implemented growth process is provided in Appendix A. The actual volume occupied by the cancer cells is computed as *V* (*t*) = *ρN*(*t*), where *ρ* = 10^−5^*mm*^3^ refers to a single cancer cell, and is estimated from the results of Del Monte [14].

According to the volume growth model, we define a size threshold, *V*_*symptoms*_, which represents the onset of symptoms in the virtual patient. In real-life scenarios, symptoms may arise at different disease stages, thus *V*_*symptoms*_ can vary across different simulations based on the clinical criteria outlined in Section 2.3. In our model, this threshold determines the start of therapy: once the cancer volume exceeds *V*_*symptoms*_, the virtual patient begins treatment according to the therapy schedule.

#### (ii) Immune interaction with cancer cells

The immune response against cancer is modeled by incorporating the interaction of cancer cells with key IS players, as observed in clinical and experimental settings, namely: TC cells, NK cells, and, to a lesser extent, antibodies (ABs) [15, 16].

Through a stochastic process, cancer cells can trigger the IS response. This activation leads to the release of cellular debris and danger signals, that, in turn, enable the IS to target cancer cells. This process is mediated through the clonal expansion of cancer-specific lymphocytes and the recruitment of non-specific activated cells, such as NK cells, at the tumor site.

Shall there be a lack of immune response, the cause is often attributed to the cancer’s ability to *evade* detection through mechanisms resembling natural selection, as described in Garrido and Aptsiauri [17]. In the absence of treatment (see next section), the progression of the disease depends on both the rate of cancer cell replication and the proportion of cells that successfully evade the IS.

#### (iii) Drug effect

The drug response processes occurring at the microscopic level -and specifically at the agent level [18]-are modeled as stochastic events, and their occurrence probabilities must be estimated from experimental data. In the simulations, the drugs effect is represented by a probability of cancer cell death, *P*. During each time step [*t, t* + Δ*t*], each cancer cell has a probability of being killed by the drug, defined as: *P* (*c*(*t*), Δ*t*) = 1 − *e*^−ν(*c*)Δ*t*^

where *c*(*t*) is the drug concentration at time *t*, and ν(*c*) is the cell-killing rate associated with that concentration. To estimate the two paramters *c*(*t*), ν(*c*), we rely on two types of experimental data: *pharmacokinetics (PK) curves*, which provide information on the concentration of the drug in tissues or fluids at different times post-administration [19], and *dose-response (DR) curves* [20], which describe the drugs effect on a target at various concentrations. Detailed calculations of drug concentration, *c*(*t*), and killing rate, ν(*c*), are provided in Appendix B.

The cell death induced by the drug releases cellular debris -similar to the response triggered by cancer cells in the IS-which then activate the immune response as described in the previous section.

This is crucial for the success of the treatment: the IS readiness to target surviving cancer cells immediately after therapy increases the likelihood of cancer clearance, as shown in Section 3.

#### (iv) Resection

We model *resection* (surgical ablation of cancer) as a stochastic process that eliminates a portion of the cancer cells that survived chemotherapy. The only effect of resection in our model is the removal of cancer cells, and we do not account for the resulting inflammatory response. The resection process is defined by the time *t*_ϵ_ of surgery and the percentage ϵ of cells excised: at *t* = *t*_ϵ_, each cancer cell has a probability *p* = ϵ of being removed, with *t*_ϵ_ and ϵ set according to literature values.

We do not consider the removal of other entities, nor the case of complete cancer cell resection, which would lead to a trivial dynamic (in our model, once all cancer cells are removed, they cannot be reintroduced), therefore, in the following simulations, some cancer cells will always remain in the system after resection.

### 2.3. Adapting the model to simulate HB

#### Cancer cells

Molecular analyses [21] have shown that the high heterogeneity of HB can be categorized into two major transcriptomic subtypes: C1, primarily associated with fetal histology, and C2, characterized by a higher percentage of highly proliferative embryonal cancer cells. Following the recent work of Failli et al. [22], this characterization will serve as the focus of our computational analysis. In fact, while additional signatures have been identified, these two remain the most extensively studied; for a comprehensive review, see [23]. The C1 subtype responds well to therapy, with 3-year overall survival (OS) and event-free survival (EFS)^1^ rates of 90 − 95% and 87%, respectively [21, 24, 25].

In contrast, the C2 subtype is associated with significantly reduced survival probabilities, with 3-year OS and EFS rates of 68% and 63%, respectively [21, 24].

To date, the mechanisms that characterize the C1 and C2 subtypes at the cellular level remain poorly understood. As a consequence, to capture the key characteristics of these subtypes, we introduced two types of cancer cells in our model: *aggressive cancer* (AC) cells and *non-aggressive cancer* (NC) cells, which differ primarily in their mean duplication times, AC cells having shorter duplication times than NC cells.

We assume that cancers of the C1 subtype are predominantly composed of NC cells, whereas the C2 subtype contains a majority of AC cells. This simplified model enables the representation of C1 and C2 subtypes in terms of the relative abundance of NC and AC cells within the tumor volume (see section 2.4).

Our ambition is to use this simple characterization -in the absence of known detailed molecular mechanisms that we could computationally model-to reproduce the key features of C1 and C2 subtypes, including their distinct proliferation behaviors and clinical outcomes.

#### Onset of symptoms and therapy administration

In HB, the *Pre-treatment Extension of Disease* system (PRETEXT, Meyers et al. [26]) is used to assess diagnosis and tumor progression (Roebuck et al. [27], Meyers et al. [26]). PRETEXT stratifies patients based on the extent of liver involvement by cancer, dividing the liver into four anatomical sections. Patients are classified into PRETEXT groups I-IV according to the number of contiguous liver sections free of cancer: I (three contiguous sections cancer-free), II (two contiguous sections cancer-free), III (one contiguous section cancer-free), and IV (no sections cancer-free).

Similarly, in our model, PRETEXT stratification is used both for evaluating disease severity and guiding treatment decisions. Initially, each virtual patient is randomly assigned a PRETEXT stage, following the proportions observed in the clinics, i.e.: 10% PRETEXT I, 34% PRETEXT II, 39% PRETEXT III, and 17% PRETEXT IV (Cairo et al. [24]).

Therapy initiation is triggered when the cancer volume reaches a stage-specific threshold, *V*_symptoms_, representing the volume at which symptoms typically appear.

We associate a threshold to each PRETEXT staging (Table 1) based on liver sectors’ dimensions (data from Mise et al. [28], Sharma et al. [29]) and irrespective of contiguity. We further assume that a sector is considered non-cancer-free if occupied by cancer cells exceeding 0.25 of its volume, approximately twice the typical detection volume.

**Table 1.** Cancer volume threshold values corresponding to the onset of symptoms, extracted from Mise et al. [28], Sharma et al. [29]. *V*_*tot*_ is the simulated total volume, chosen to contain a maximum of 10^5^ cancer cells.

| PRETEXT group | $V_{symptoms}$ |
| --- | --- |
| PRETEXT I | $V_I = 0.0575V_{tot}$ |
| PRETEXT II | $V_{II} = 0.1055V_{tot}$ |
| PRETEXT III | $V_{III} = 0.1410V_{tot}$ |
| PRETEXT IV | $V_{IV} = 0.2460V_{tot}$ |

Importantly, as PRETEXT class is assigned to the virtual patient before simulation starts, there could be symptomatic patients with lower cancer volumes than asymptomatic ones, and vice versa. For instance, a patient with a relatively low cancer volume (e.g., *V* (*t*) = 0.1*V*_*tot*_) classified *a priori* as PRETEXT I will be symptomatic, while another with a higher volume (e.g., *V* (*t*) = 0.2*V*_*tot*_) but classified as PRETEXT IV will be asymptomatic.

To simulate the drug effect, we refer to cisplatin, the gold standard treatment for HB, and use PK and DR curves available from the literature (Peng et al. [30], Andersson et al. [31]) and from recent *in vitro* experiments (Demir et al. [32]), respectively. Note that, although different effects of chemotherapeutic drugs, and of cisplatin in particular, have been observed depending on the proliferation status of osteosarcoma cancer cells (Granada et al. [33]), there is no specific information on the effect of cisplatin on the different cells that characterize the two HB transcriptomic subtypes C1 and C2. Therefore, we assume that the interaction of the drug with NC and AC cells is the same.

Recent studies [34] indicate that chemotherapy, particularly with cisplatin (the gold standard in the treatment of HB), stimulates an anti-cancer immune response. The treatment protocol is obtained from the literature (Perilongo et al. [35]) and consists of four cisplatin-based chemotherapy administrations followed by resection and two additional chemotherapy administrations, Table 2. In our model, therefore, the drug not only reduces the cancer burden directly but also boosts the IS response and memory.

**Table 2.**
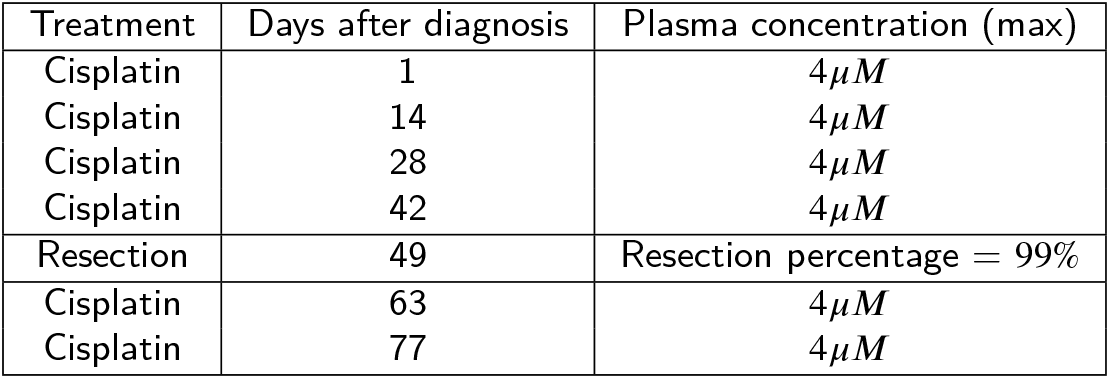
Details of the administered therapy, as obtained from Perilongo et al. [35].

### 2.4. Model parameters calibration

To the best of our knowledge, there is no available microscopic data on the interaction between HB cells and immune cells that could directly inform our model. However, evidence of such interactions exists in the literature [36, 37, 38].

Therefore, we calibrated the model using macroscopic clinical proxies, including OS and EFS rates and the distribution of the main molecular cancer subtypes C1 and C2 at the time of diagnosis. Additionally, we incorporated experimental *in vitro* data on single-cell duplicating times [32].

To facilitate comparison with clinical data, we classified simulation outcomes into four distinct categories. An outcome is classified as *Fatal* if the tumor reaches the critical threshold volume for death. *Progressive* refers to patients whose tumor continues to grow after diagnosis, with the tumor growth surpassing the detection threshold. Patients who completely eliminate cancer cells are categorized as *Cleared*, and the remaining cases are classified as *Non-progressive*. Further, patients who fall into the Fatal and Progressive groups are classified as *Symptomatic*, while Non-progressive and Cleared cases are labeled as *Asymptomatic*.

Simulation outcomes are categorized over time based on the relative tumor volume post-diagnosis (*t*_*d*_), using the following thresholds:

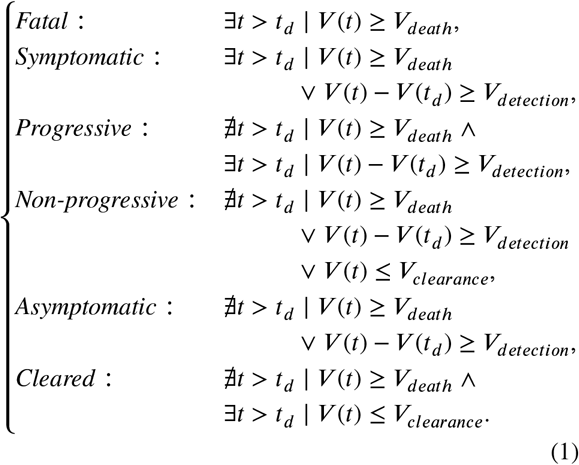

The detection threshold was set to *V*_detection_ = 7.5 × 10^−3^*V*_tot_ based on [39, 40]. The clearance threshold was set to *V*_clearance_ = 0, indicating complete tumor elimination. The death threshold was defined as *V*_death_ = 0.8*V*_tot_, based on the assumption that exceeding 80% of the total organ volume results in irreversible organ failure.

#### Calibration for Untreated Virtual Patients

Each set of parameters was tested by running 1000 simulations over a virtual period of three years. Due to the stochastic nature of the model, this process generates 1000 different virtual patients. For each patient, we recorded the temporal evolution of the cancer cell populations, including both NC and AC cells, as well as the *Molecules* and *Cells* introduced in Section 2.1.

The simulations started with an initial cancer cell population such that *V* (*t* = 0) > *V*_detection_, where *V*_detection_ represents the minimum detectable tumor volume [39, 40].

In the absence of treatment, the parameters governing cancer growth and immune interactions were adjusted to ensure that the simulated tumor growth and survival rates closely mimic the observed clinical trends. Specifically, we considered the initial number of cancer cells, the AC/NC cell ratio, and the duplication times *μ*_*AC*_ and *μ*_*NC*_. In our model, these values are linked to the C1 and C2 cancer subtypes, under the assumption that a relatively higher abundance of AC cells corresponds to a C2 classification. Data from *in vitro* experiments on the C2 subtype [32] were used to model the duplication time of AC cells as a Gaussian distribution with mean μ_*AC*_ = 2.9 days and variance *σ*^2^ = 0.5 days^2^, ensuring that AC cells duplication times fell within the range of 2.1 to 3.75 days with a probability greater than 0.95.

Similarly, NC cell duplication times were modeled as a Gaussian distribution with variance *σ*^2^ = 0.5 days^2^ and mean *μ*_*NC*_, constrained by *μ*_*NC*_ < *μ*_*AC*_. The calibration of the initial number of cancer cells, the AC/NC ratio, and *μ*_*NC*_ was performed by comparing the simulated disease outcomes, categorized as described in the previous section, to the clinical data presented in Table 3.

**Table 3.** Model capacity to simulate clinical observables. First column: clinical observable. Second column: simulated value. Third column: clinical value as extracted or computed from [24]. Specifically, the death-to-symptomatic ratio is obtained from clinical data on OS and EFS percentages according to the relation Death/Symptomatic = (1-OS%)/(1-EFS%), as discussed in the main text.

| Clinical observable | Simulated value (%) | Clinical Value (%) |
| --- | --- | --- |
| PRETEXT I - III 3 yrs Death/Symptomatic | 0.49 | 0.4 |
| PRETEXT IV 3 yrs Death/Symptomatic | 0.64 | 0.68 |
| C1 subtype 3 yrs Death/Symptomatic | 0.37 | 0.38 |
| C2 subtype 3 yrs Death/Symptomatic | 0.89 | 0.86 |
| PRETEXT I subtypes C1, C2 | 0.73, 0.27 | 0.65, 0.35 |
| PRETEXT II subtypes C1, C2 | 0.70, 0.30 | 0.72, 0.28 |
| PRETEXT III subtypes C1, C2 | 0.67, 0.33 | 0.6, 0.4 |
| PRETEXT IV subtypes C1, C2 | 0.60, 0.40 | 0.57, 0.43 |

Although current clinical data do not allow for precise quantification of the AC/NC ratio, the calibration process reveals that, to achieve qualitative agreement with real-world scenarios, C1 subtypes must contain fewer than 30% AC cells, with C2 subtypes exceeding this threshold. Furthermore, considering the differing growth rates of the two cell types, the initial proportions are randomly assigned such that, on average, at time *t* = 0, there is one AC cell for every 10^3^ NC cells. This configuration ensures that the cell population dynamics reproduce a logistic growth in line with the decreasing relative growth rates observed in human cancers [41, 42].

#### Calibration for Treated Virtual Patients

After calibrating cancer growth and immune interactions for untreated patients, we introduced therapy effects. Since treatment calibration may require adjusting parameters that were previously validated for untreated patients, an iterative process was necessary to ensure consistency.

Therapy parameters were selected to closely match experimental values, minimizing the difference between simulated outcomes and clinical data. Specifically, the 3-year OS and EFS rates can provide constraints that the simulation outputs need to satisfy. Given a population of *m* individuals, the number of survivors, dead, asymptomatic, and symptomatic patients can be calculated as *mOS*%, *m*(1− *OS%*) *mEFS*% and *m*(1 − *EFS*%), respectively. However, these relationships cannot be used to compare the simulation results with clinical observations, as the latter are conducted on a population that includes individuals who have cleared the tumor through complete resection, a scenario not allowed by our model.

We therefore defined:

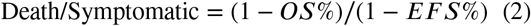

where the numerator accounts for the number of *Fatal* outcomes and the denominator accounts for the number of *Fatal* plus *Progressive* outcomes. This quantity guarantees the alignment of the simulated and clinical outcome groups, while excluding complete surgical resection from the model, thereby reducing the overall computational burden.

The first column of Table 3 categorizes the calibration data into two main groups: (i) clinical percentages at 3 years in symptomatic cases (first four rows) and (ii) clinical percentages of the two major cancer subtypes across different PRETEXT groups at resection (second four rows). The values for the first category were derived from clinical OS and EFS percentages using Equation 2.

## 3. Results

### 3.1. ABM for clinical outcome simulation

Once the best configuration of parameters has been identified through the calibration process described in Section 2.4, we created a new cohort of 5000 HB virtual patients over three years time. As expected, untreated virtual patients face fatal outcome (Figure 1 and first column of Table 4), in agreement with what was observed in the pre-chemotherapy era [43, 44]. Note that the tumor growth is well approximated by a logistic curve with a growth rate of 0.025 days. It is important to note that, even in the case of fatal outcome, a difference in dynamics can be observed, particularly regarding the time it takes for cancer to reach the *V*_*death*_, as it can be seen in the divergence from the mean trend shown in Figure 1. These differences are due both to the stochasticity of individual processes occurring during simulations, and to intrinsic differences in the IS of virtual patients.

**Table 4.**
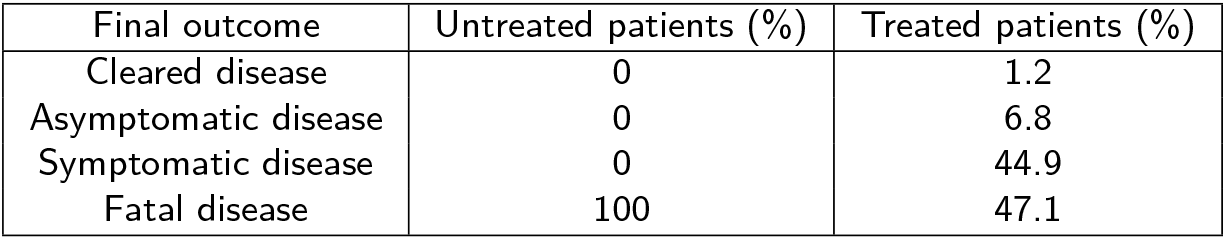
Simulated disease outcome. First column: final outcome of the simulated disease. Second column: percentage of untreated virtual patients per outcome class. Third column: percentage of treated virtual patients per outcome class.

| Final outcome | Untreated patients (%) | Treated patients (%) |
| --- | --- | --- |
| Cleared disease | 0 | 1.2 |
| Asymptomatic disease | 0 | 6.8 |
| Symptomatic disease | 0 | 44.9 |
| Fatal disease | 100 | 47.1 |

**Figure 1:**
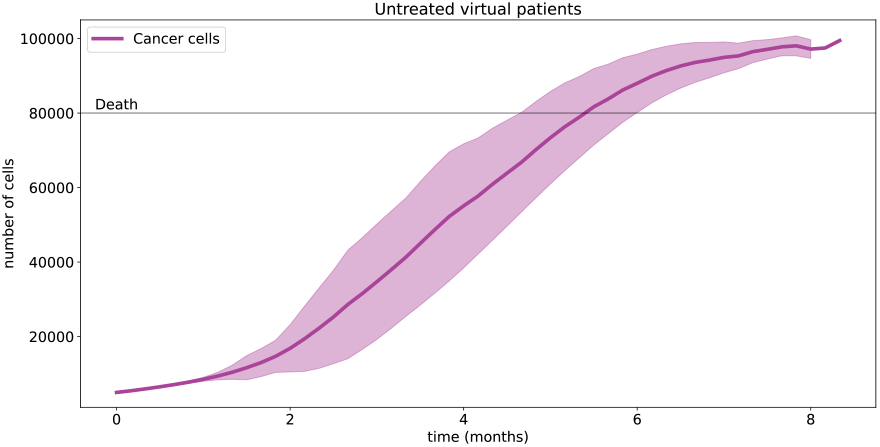
Cancer evolution in untreated virtual patients. On the *x*-axis: time (months). On the *y*-axis: number of cancer cells per *mm*^3^ (median value and interquartile range). Magenta solid line: total number of cancer cells.

We repeat the simulations carried out in the case of untreated patients, all parameters being equal, but administering standard therapy. Therapy begins when the simulated cancer volume reaches the threshold for the onset of symptoms, as discussed in Section 2.3, and we classify the disease outcome according to the percentage of volume occupied by cancer cells, as described in Section 2.4.

When patients undergo therapy, the whole range of outcomes is possible as shown in the second column of Table 4, and in particular patients whose tumor mass is completely cleared (Figure 2 panel a); constrained below the threshold of symptom onset (Figure 2 panel b),; leading to non-fatal disease progression (Figure 2 panel c), and fatal despite treatment (Figure 2 panel d).

**Figure 2:**
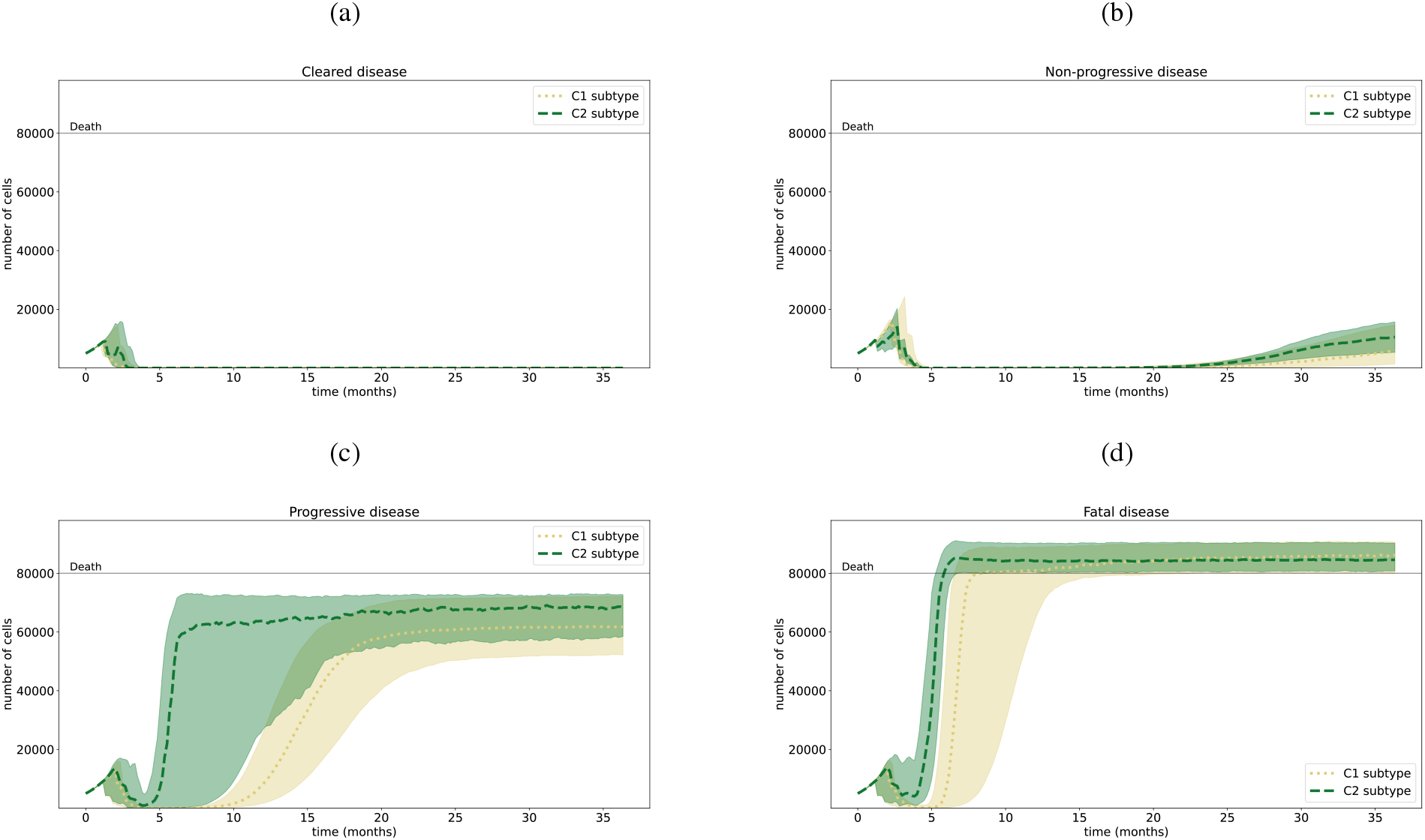
Cancer evolution in treated virtual patients. On the *x*-axis: time (months). On the *y*-axis: number of cancer cells per *mm*^3^ (median value and interquartile range). Yellow dotted line: HB C1 subtype. Green dashed line: HB C2 subtype. Panel a: cleared disease. Panel b: non-progressive disease. Panel c: progressive disease. Panel d: fatal disease.

Since complete resection is not allowed in our simulations, the clearance is due to the combined effect of drug therapy and IS.

We can observe that the simulated reduction in cancer size during therapy is in agreement with that measured in clinical cases for HB: the ratio between the mean volume after two cycles of chemotherapy and the mean baseline volume, as well as the ratio between the mean volume before resection and the mean baseline volume, are 0.42 ± 1.43 and 0.31±1.06 in the simulations, and 0.55±0.75 and 0.45±1.24 in real patients, respectively, with clinical values obtained from Nguyen et al. [45].

Moreover, our model correctly reproduces the percentage of patients exhibiting a C1 or C2 cancer subtypes at the time of resection [24] and the ratio of dead patients to symptomatic patients three years after diagnosis. Furthermore, the ratio of dead to symptomatic patients is also correct for individual cancer subtypes, confirming the consistency of the model, Table 3.

Finally, we note that virtual patients with the same outcome may show different disease progressions. In particular, we observe that different evolutions of the number of NC and AC cells can lead to the same macroscopic behaviour. Figure 3 focuses on how different populations of NC and AC cells all lead to fatal disease. Similar results were found for the other categories (data not shown).

**Figure 3:**
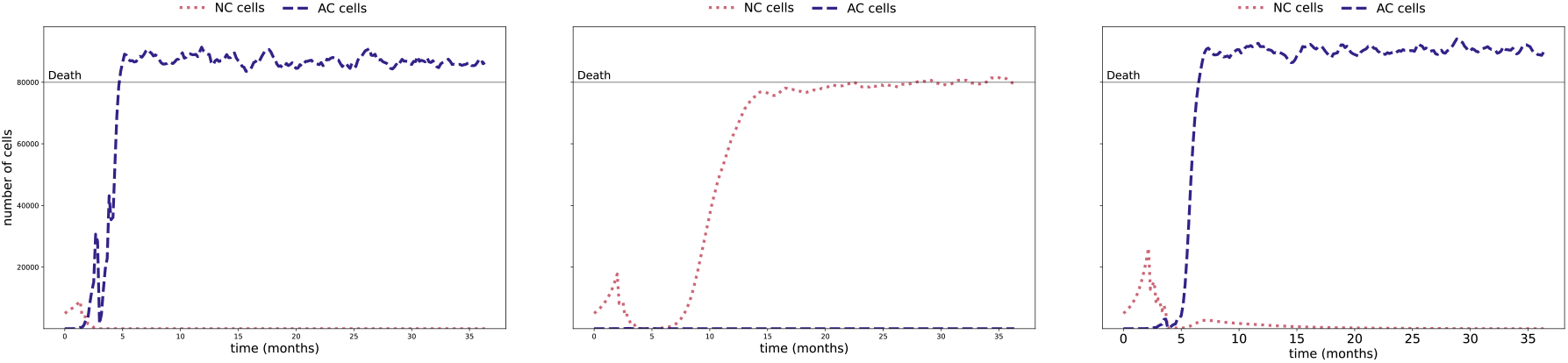
Cancer evolution in treated virtual patients with fatal disease: detail of the different dynamics. On the *x*-axis: time (months). On the *y*-axis: number of cancer cells per *mm*^3^. Red dotted line: number of NC cells. Blue dashed line: number of AC cells. Left panel: presence of AC cells only in the long term. Central panel: presence of NC cells only in the long term. Right panel: majority presence of AC cells in the long term, majority of NC cells during treatment.

### 3.2. ABM for microscopic level insight into clinical outcome

We then turned to the immunological entities modeled to explore potential correlations with clinical outcomes, specifically by analyzing their values at clinically relevant time points, namely at the start of therapy, the day of resection, the end of therapy, and one month after the end of therapy (follow-up). To quantify this correlation, we introduce a relative difference (*RD*) computed as:

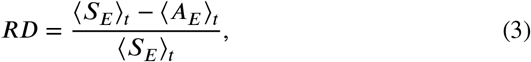

where *E* is the immunological entity of interest, *t* is the clinically relevant time point introduced above, and ⟨*S*_*E*_⟩_*t*_ and ⟨*A*_*E*_⟩_*t*_ are the mean values of *E* computed at time *t* for the symptomatic and asymptomatic virtual patients, respectively. The farther the normalized difference for an immunological entity is from 0, the more relevant that entity may be for discriminating between symptomatic and asymptomatic patients.

In Figure 4, we display only the entities with *RD* standard deviation smaller than the mean value, i.e. entities whose variation is sufficiently stable across measurements. At the start of therapy, no significant difference could be observed between the values of immunological entities in symptomatic and asymptomatic patients (data not shown), confirming that, without the stimulation resulting from drug therapy, the IS is unable to mount an adaptive response against HB.

**Figure 4:**
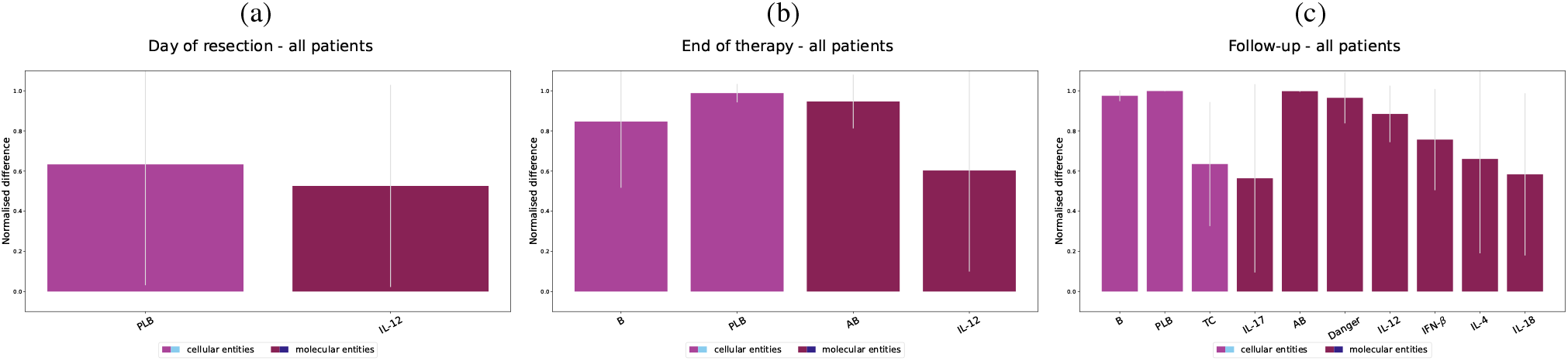
Correlation between immunological entities and disease in virtual patients. On the *x*-axis: immunological entities. On the *y*-axis: relative difference, as defined in Equation 3. Magenta bar: cellular entities. Purple bar: molecular entities. Panel a, b, c: results for all virtual patients on the day of resection, end of therapy, and day of follow-up, respectively.

On the other hand, other time points show that B, PLB and ties TC cells are relevant cellular entities. In fact, these entigrow in number when they encounter and recognise an antigen, and are therefore an indicator of the death of cancer cells and the subsequent release of the associated antigen into the system. We also observe a significant discrepancy, especially on the day of follow-up, for cytokines IL-12, IL-18, IFN-*β*, IL-4 and IL-17. The two former have already been observed in association to HB: cytokine IL-12, in particular, plays a major role in triggering anti-cancer immune responses [46, 47], while both IL-18 and IL-12 levels were observed to be high in HB patients, with the serum IL-18 supposed to be an independent prognostic survival factor for patients with liver cancer [48, 49]. High levels of expression of the cytokines IL-4 and IL-17, on the other hand, have been observed in the liver cancer Hepatocellular Carcinoma [50], while IFN-*β* has shown to play an important role in controlling the same tumor [51].

These results are in agreement with a scenario in which immunological variables measurable at clinically accessible time may constitute a risk factor for HB, offering the potential to enhance the monitoring of immunological variables to design appropriate therapeutic measures.

## 4. Discussion and summary

In this work, we use ABM C-IMMSIM to study the combined effect of immune response and therapy against HB to follow the dynamics of immunological variables and model possible disease outcomes. To this end, we introduced cancer cells with different growth features, allowing us to reproduce the macroscopic growth of HB and the behavior of HB subtypes and a therapy model based on experimental data.

Our simulations allow us to reproduce relevant observed clinical data, in particular the ratio between the 3-years OS and EFS percentages, expanding the relevance of C-IMMSIM to tumor contexts. In particular the granularity of the model is capable of reproducing heterogeneity, a determinant feature of cancers in general and of HB in particular, owing to the capability to reproduce observations made on the main C1 and C2 subtypes, suggesting that the model is consistent and usable for analyzing the interaction between immunologic variables and disease outcome. Moreover, based on an analysis of the simulation results at the microscopic level, a series of potentially relevant markers is detected, including B cells, cytotoxic T lymphocytes, and the cytokines IL-12, IL-18, IFN-*β*, IL-4 and IL-17. Further validation of the clinical potential of immunological quantities as therapeutic markers represent potential future developments.

### A. Cancer growth model

We assume that the growth rate *μ* of each cell is constant over each simulation time step:

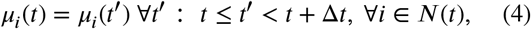

where *N*(*t*) denotes the population of cancer cells present in the system at time *t*. Moreover, we assume that each cell *i* is characterised by a duplication rate 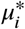 that is inherited by its offspring. This latter condition allow^*i*^ us to consider the replication process of each cell during the simulation time step to be a simple birth, or Yule, process [52].

The Yule process describes the number *N*(*t*) of individuals of a population at time *t* starting from the number *N*(0) = *n*_0_ of individuals at time *t* = 0 assuming a constant growth rate *μ* equal for each individual in the population. In particular, the probability distribution of the number of individuals *N*(*t*) for the Yule process is described by the following expression:

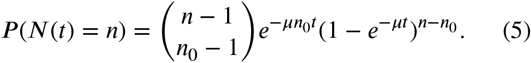

In C-IMMSIMm we use the Yule process with *n*_0_= 1 to estimate the offspring of each cancer cells generated during the simulation time step Δ*t*. Note that the actual growth rate *μ*_*i*_ of the *i*-th cell varies from one simulation step to the next depending on the state of the system (Equation 4), i.e.,, we perform different Yule processes at each time steps for each cancer cell.

Let a site be a lattice point and *V*_*s*_, *V*_*c*_, *K* = *ηV*_*s*_/*V*_*c*_ the volume of the site, the volume of a cancer cell and the carrying capacity of the site (i.e., the maximum size of the cell population within the site), respectitely. The constant *η* > 0 takes into account resource competition between cancer cells. Moreoter, let *r*_*s*_ and *r*_*c*_ < *r*_*s*_ be the radius of sphere of volume *V*_*s*_ and of a cancer cell, respectively. We define *V*_*e*_ as the volume of a spherical shell identified by the difference between the sphere of radius *r*_*s*_ and the sphere of radius *r*_*s*_ − *r*_*c*_. During the cancer growth processes, the *i*-th cell placed in the *site x* is assumed to be in the shell of the *site* with probability *q* = *V*_*e*_/*V*_*s*_ and in the center of the *site* with probability 1 − *q*. If the cell is in the center of *x*, its offspring is generated in that *site*. If the cell is in the shell, its offspring is generated in *x* with probability *p* = 1 − [*N*_*x*_(*t*)*V*_*c*_/*V*_*s*_], otherwise a new *site* is chosen at random from the neighborhood of *x*. Let *y* ∈ {*x*, neighboring of *x*} be the *site* where the offspring will be placed. During the time interval (*t, t* + Δ*t*), the number of cells *N*_*i*_ generated from the *i*-th cell is obtained from Equation 5 with growth rate 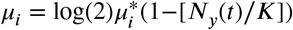.Although this model may not be properly descriptive of the phenomenon of growth of a cancer mass, it allows its phenomenology to be reproduced. In particular, the resulting cancer growth follows a logistic curve, Figure 1, that is the most appropriate modelling of the process according to experimental data [53, 42].

### B. Introducing drug therapy

#### B.1 Including pharmacokinetics

Common cancer chemotherapy treatments consist of several drug doses. After each administration the drug concentration peaks and then decays due to absorption and elimination. As a result, the drug concentration cannot be considered constant during the therapeutic treatment. In addition, the drug ability to reach the target cells depends on the tissue where the cells are located [19]. Therefore, PK must be included in the analysis.

PK is a branch of pharmacology that aims to study the evolution of drugs administered to a living organism over time [19, 54]. Different mathematical models can be used [55], here, we include a simple model consisting of a single compartment. This simplified model allows us to perform an extended analysis of the therapy outcome as a function of the other model’s parameters.

As a first approximation, we assume that the concentration *c*_*n*_(*t*) of the drug reaching the *n*-th target cells is spatially homogeneous, and it can be written with a simple proportional formula:

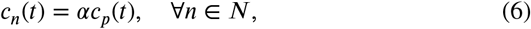

where *N* is the set of cancer cells, α ≥ 0 is a proportionality constant estimated from the literature and *c*_*p*_(*t*) is the *simulated plasma concentration* i.e., the step-like function that approximates the experimental concentration curve by a series of daily, constant, values (see Figure 5).

**Figure 5:**
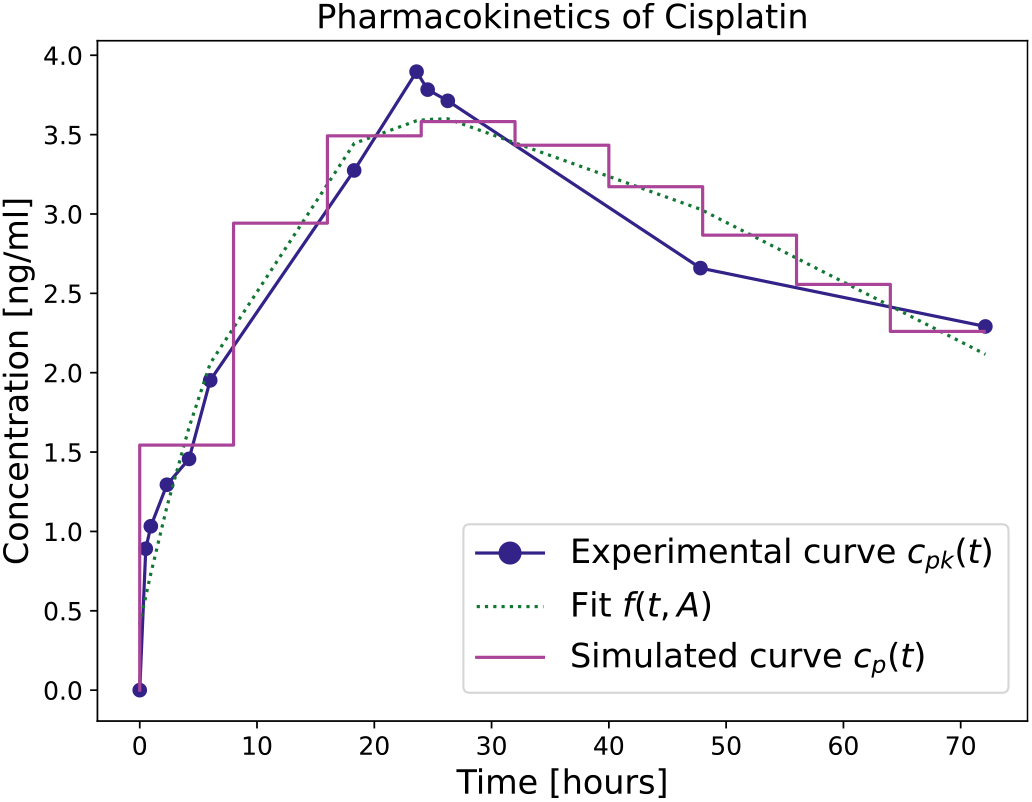
PK curves of cisplatin. Blue points: experimental points of plasma concentration over time, obtained from Peng et al. [30]. Green dotted line: fit of experimental points with function *f* (*t*, **A**) (Equation 8). Magenta solid line: simulated plasma concentration *c*_*p*_(*t*) (Equation 10).

Under these assumptions, we can evaluate the amount of drug reaching cancer cells starting from the experimental PK curve *c*_*pk*_(*t*) describing the plasma concentration of drug over time as follows: let *f* (*t*, **A**) be an analytic function whose parameters **A** are obtained by fitting the experimental curve *c*_*pk*_(*t*) with *f* (*t*, **A**). The *simulated plasma concentration* 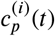 of drug at time *t* = *d*_i_ + *m*Δ*t* due to the administration of drug on day *d* is computed as:

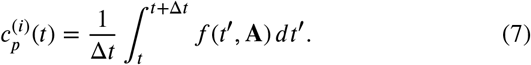

In particular, we only consider the following expression for function *f* (*t*, **A**):

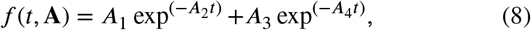

with the additional constraint that the drug concentration is zero one week after the administration. The total *simulated plasma concentration* of drug *c*_*p*_(*t*) resulting from *M* administrations on days *d*_1_, …, *d*_*M*_, *d*_*M*_ ≤ *t*, is the following:

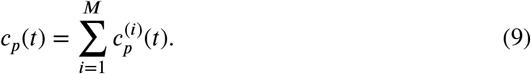

By substitution, we obtain the concentration *c*(*t*) of drug interacting with each administered cancer cell:

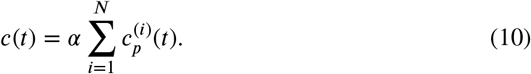

Figure 5 shows the experimental curve *c*_*pk*_(*t*) and the resulting simulated plasma concentration 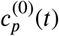 in case of a single day administration of drug at *d*_0_ = 0

#### B.2. Estimating drug-induced death probability

A possible approach to model the drug effect in the ABM is the introduction of a probability *P* for the drug-induced cell death during the time interval Δ*t*. We assume that:

- the drug concentration is homogeneous and constant within the time step Δ*t*;
- the rate ν of drug-induced death depends only on the drug concentration;
- the drug-induced cell death is a stochastic memoryless process.

Therefore, the probability ***P*** ≡ ***P*** (*c*, Δ*t*) can be written as:

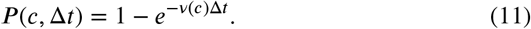

The death rate ν can be estimated from the experimental DR curves, that describe the response *R*(*c, T*) of a treated cell population as a function of the concentration *c* of the administered drug and the duration *T* of the treatment. The DR curves are obtained from controlled experiments where target cells, kept in a closed system, are treated with a desired dose of drug that is assumed to be constant during the experiment. In particular, *R*(*c, T*) can be defined as the relative cell count [20, 56, 57]:

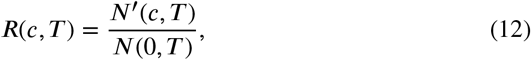

where *N*(*c, T*)^′^ and *N*(0, *T*) are the number of treated and control (untreated) cells measured at the end of the experiment of duration *T*, respectively. In order to estimate ν(*c*) from the DR curves, we assume that the growth rate *μ* and the death rate ν(*c*) are the same for each cell and constant in time during the experiment aimed at evaluating the DR curve. It follows that the evolution over time of the number of treated and control cells can be written as:

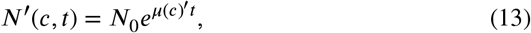

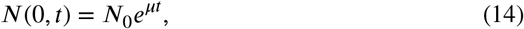

where *N*_0_ is the number of cells at the beginning of the treatment and *μ*(*c*)^′^ = *μ* − ν(*c*).

Note that, in general, the death rate ν (and thus the growth rate *μ*′) depends on the concentration *c* of the administered drug, that is, ν ≡ ν(*c*), *μ*′ ≡ *μ*′(*c*). By substituting Equations 13 and 14 in Equation 12, we obtain:

Equations 13 and 14 in Equation 12, we obtain:

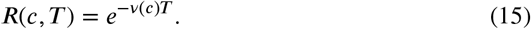

Therefore, by knowing the experimental DR curve *R*(*c, T*), the drug concentration-dependent death rate ν(*c*) can be estimated as:

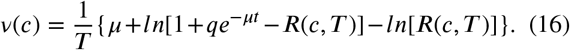

In particular, *R*(*c, T*) can be approximated by the function *g*(*c*, **B**) [58]:

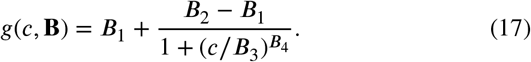

We obtain the parameters **B** by fitting the experimental curve *R*(*c, T*) obtained from the administration of cisplatin to the hepatoblastoma cell-line XT243 [32] with the function *g* (*c*, **B**), in order to evaluate ν (*c*). The DR curve and the Resulting values of *P* (*c*, Δ*t*) are shown in Figure 6. We use the experimental data for cell-line XT243 as a reference value to estimate the effect of the drug on the tumour cells in our *in silico* model.

**Figure 6:**
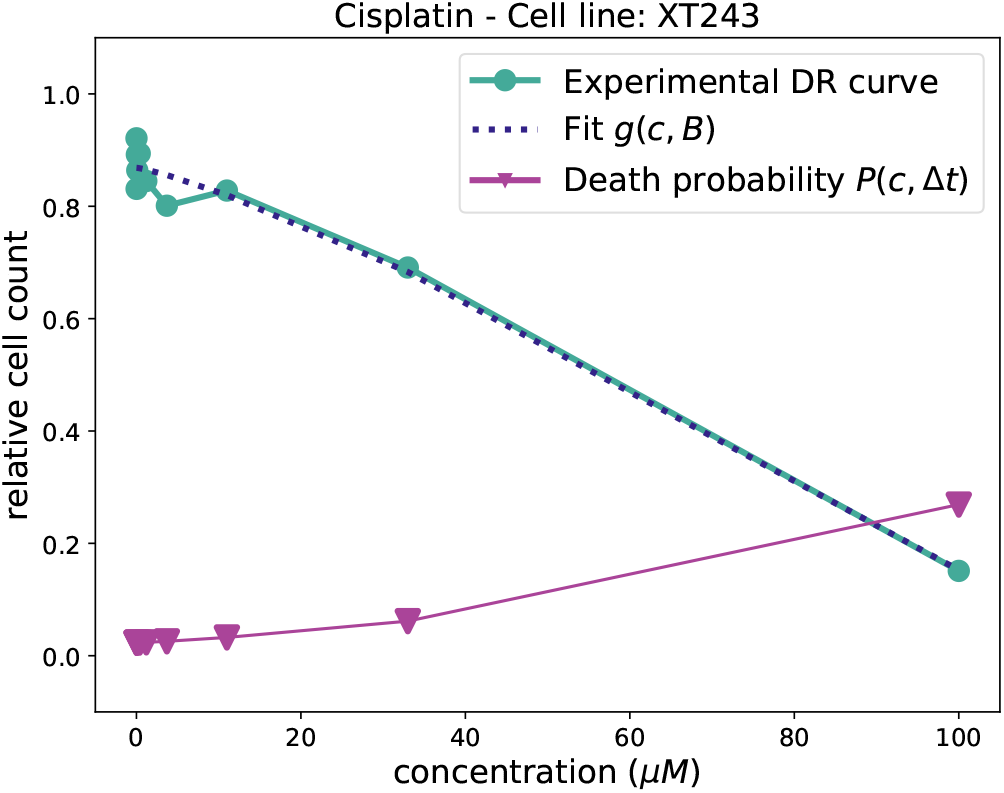
DR curves of cisplatin. Blue points: experimental points of DR curve obtained from the administration of cisplatin to the hepatoblastoma cell-line XT243, data from Demir et al. [32]. Green dotted line: fit of experimental points with function *g*(*c*, **B**) (Equation 17). Magenta triangles: estimated death probability *P* (*c*, Δ*t*) (Equation 11).

Note that other cell growth models can be considered to estimate *N*′(*c, T*) and *N*(0, *T*) [59], resulting in a different expression of the function ν(*c*).

For the sake of completeness, we remark that Equation 11 can also be obtained by defining the death probability *P* (*c*, Δ*t*) as the ratio between the number of cells 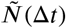 killed by the drug at concentration *c* and the number of cells which the drug can act upon during the time interval [*t, t* + Δ*t*]. In particular, by assuming that both the growth rate *μ* and the death rate ν are constant in time, we obtain:

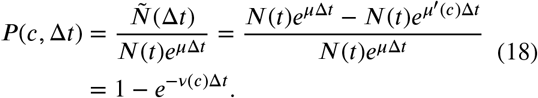

## Data Availability

All data produced in the present study are available upon reasonable request to the authors

## Acknowledgments

The authors would like to thank Jean-Fançois Moreau for his careful feedback.

## Footnotes

1 OS is defined as the time from diagnosis to death or censoring at the time of last contact. EFS is defined as the time from diagnosis to the first occurrence of disease progression, death, or censoring at the time of last contact.

